# ENCoDE – a skin tone and clinical dataset from a prospective trial on acute care patients

**DOI:** 10.1101/2024.08.07.24311623

**Authors:** Sicheng Hao, Joao Matos, Katelyn Dempsey, Mahmoud Alwakeel, Jared Houghtaling, Chuan Hong, Judy Gichoya, Warren Kibbe, Michael Pencina, Christopher E. Cox, A. Ian Wong

## Abstract

**Background:** Although hypothesized to be the root cause of the pulse oximetry disparities, skin tone and its use for improving medical therapies have yet to be extensively studied. Studies previously used self-reported race as a proxy variable for skin tone. However, this approach cannot account for skin tone variability within race groups and also risks the potential to be confounded by other non-biological factors when modeling data. Therefore, to better evaluate health disparities associated with pulse oximetry, this study aimed to create a unique baseline dataset that included skin tone and electronic health record (EHR) data.

**Methods:** Patients admitted to Duke University Hospital were eligible if they had at least one pulse oximetry value recorded within 5 minutes before an arterial blood gas (ABG) value. We collected skin tone data at 16 different body locations using multiple devices, including administered visual scales, colorimetric, spectrophotometric, and photography via mobile phone cameras. All patients’ data were linked in Duke’s Protected Analytics Computational Environment (PACE), converted into a common data model, and then de-identified before publication in PhysioNet.

**Results:** Skin tone data were collected from 128 patients. We assessed 167 features per skin location on each patient. We also collected over 2000 images from mobile phones measured in the same controlled environment. Skin tone data are linked with patients’ EHR data, such as laboratory data, vital sign recordings, and demographic information.

**Conclusions:** Measuring different aspects of skin tone for each of the sixteen body locations and linking them with patients’ EHR data could assist in the development of a more equitable AI model to combat disparities in healthcare associated with skin tone. A common data model format enables easy data federation with similar data from other sources, facilitating multicenter research on skin tone in healthcare.

**Description:** A prospectively collected EHR-linked skin tone measurements database in a common data model with emphasis on pulse oximetry disparities.

## Introduction

Pulse oximetry, a fundamental non-invasive tool used in healthcare settings worldwide, enables the monitoring of oxygen saturation levels and heart rate through the use of light-emitting diodes at specific wavelengths (660 nm and 940 nm).^22^ This technique, by measuring differences in light absorption by oxyhemoglobin and deoxyhemoglobin, estimates the peripheral oxygen saturation (SpO_2_) as a surrogate for arterial oxygen saturation (SaO_2_), providing critical data for patient triage, monitoring, and intervention.^2^ Despite its widespread use and the immediacy of its results, pulse oximetry’s reliability is increasingly questioned, especially in critical care settings where accurate measurements are vital.^3^ The device’s fundamental dependence on optical technology makes it susceptible to various factors that can skew readings, such as patient movement, ambient light interference, and, most critically, variations in skin pigmentation.^4–10^

Recent studies highlight significant racial and ethnic discrepancies in SpO_2_ readings, which could obscure the true severity of conditions such as hypoxemia in individuals with darker skin tones.^4,15^ These inaccuracies in pulse oximetry can lead to mismanagement of critically ill patients, resulting in severe consequences, including organ dysfunction and death.^11^ Furthermore, these discrepancies often result in delayed or inadequate interventions, exacerbating health inequities and complicating clinical decision-making.^4–10^ Studies also suggest that conventional calibration methods for pulse oximeters may not account for physiological variations across different skin tones, underscoring the need for tailored approaches.^12,13,23^ The underlying issue has been traced to differences in skin tone, which affect the oximeter’s light absorption readings. Yet, the integration of skin tone as a medical variable has been limited, leading to a substantial knowledge gap in how pulse oximetry should be adjusted or interpreted based on this factor.^8^

To bridge this gap, our research, encapsulated in the ENCoDE (mEasuring skiN Color to correct pulse Oximetry DisparitiEs) study, aims to systematically collect and analyze skin tone data at different body locations from patients requiring acute care. This study is designed to develop a robust framework for incorporating skin tone into clinical assessments, ensuring that pulse oximetry can provide accurate readings across a diverse patient population. By linking these findings with clinical data in electronic health records (EHR), we hypothesize that our approach will significantly reduce the disparities currently observed in pulse oximetry, thereby enhancing patient care and outcomes.

## Methods

### IRB

This study was approved by the Duke Health IRB under Pro00110842 on 18 May 2022, titled “ENCODE (mEasuring skiN Color to correct pulse Oximetry DisparitiEs).”

### Cohort Acquisition

The ENCoDE project enrolled patients admitted to inpatient care at Duke University Hospital (Durham, NC, USA) with hospital encounters. The requirement for inclusion was synchronized ABG-pulse oximetry measurements, defined as at least one pulse oximetry value recorded within 5 minutes prior to an ABG value captured in EHR and referred to as SaO_2_-SpO_2_ pairs. All data, including patient consent, measurements, and EHR data, were stored in REDCap electronic data capture tools hosted at Duke University.^24,25^ For patients unable to consent, a Legally authorized representative (LAR) would consent on their behalf, and the patient re-consented after they regain the ability to consent. Exclusion criteria included unremovable fingernail polish, admission for vascular complications (e.g., grafting or stenting), any limb amputations, and causes of skin discoloration such as vitiligo, jaundice, and wounds/bruising. These criteria were established to ensure data quality by avoiding cases of arterial insufficiency or other conditions that could affect skin tone across all patient locations.

### Data Collection

#### Skin tone data collection

In all patients, four modalities of skin assessments were conducted: infrared temperature (using the HoMedics HTD8813C [clinical-range, 34-42.9 C°] and IDEAL Model #61-847 [general-range, -32 - 500 C°]), administered visual scales (Fitzpatrick Skin Type, Monk Skin Tone, and Von Luschan), colorimetric (Delfin SkinColorCatch), spectrophotometric (Konica-Minolta CM-700d, Variable Spectro 1 Pro), and photography via mobile phone cameras (Google Pixel 4a, iPhone SE 2020). Visual skin scales were printed for reference on 4“x 6” photo paper.

Measurements were taken using all four skin assessment modalities at sixteen different locations: eight on the left and right upper extremities (dorsal and ventral finger pad, dorsal and ventral palm), three on the head (forehead, inner and outer surface of an earlobe), one on the sternum, and four on the left and right lower extremities (dorsal and ventral toe). Measurements were collected from patients lying down or in a seated position. A black card was placed on the opposite side for earlobes (and fingers, if needed) to reduce the impact of reflection. For this study, measurements were collected among patients admitted to an intensive care unit (ICU) and regular nursing floor units.

The study utilized two trained personnel to collect measurements to improve the timeliness of data collection and create an efficient workflow in a dynamic, multidisciplinary, and fast-paced environment. The location of the pulse oximeter was reported as directly observed by the clinical research coordinator at the time of data collection or surveyed by clinical staff regarding location at the time of the ABG.

#### EHR data

Patients’ hospital encounter information, demographic details, laboratory measurements (such as arterial blood gas panel, complete blood count panel, and comprehensive metabolic panel), and flowsheets (containing measurements of standard vital signs and information about oxygen delivery) were extracted from Duke University Hospital’s EHR system (EPIC Clarity). All data, including image records, are linked to the patient’s encounter using unique hospital account identifiers.

### Data Processing

#### Data linking

To link skin tone data with patients’ EHR data, we pulled all structured data from EPIC Clarity and REDcap into Duke’s Protected Analytics Computing Environment (PACE) for processing. Tables from the two systems are linked via the hospital encounter number and the patient’s medical record number (MRN).

#### Image feature processing

Every image captured by mobile devices undergoes a filtering process to isolate the brightest section, achieved by selecting the largest contour above the median brightness in greyscale. Subsequently, a mask is generated to eliminate extraneous values. An example processed image, with full consent from the measurement subject, is provided below (Figure 2). From these masked images, the average and standard deviation on each RGB (Red, Green, and Blue) and LCH (Lightness, Chroma, and Hue) channels are extracted as features linked to the patient’s skin tone.

#### De-identification

We de-identified our dataset according to the provision of the Health Insurance Portability and Accountability Act (HIPAA) Safe Harbor. All date and time information has been shifted randomly into the future while preserving the difference within one subject. Patient encounter numbers and MRNs are randomly re-mapped to visit_occurrence_id and patient_id. All measurement values are first evaluated as strings and then whitelisted before deidentification. Image data from potential biometric locations (ventral side of both left and right fingers, palms, and toes) are excluded. However, processed features (e.g., mean or standard deviation RGB values) from images that are not considered biometric information are included.

#### OMOP Conversion

The Observation Medical Outcomes Partnership (OMOP) Common Data Model (CDM) is a standardized framework designed to enable systematic analysis of disparate observational healthcare databases, facilitating large-scale data integration and research. We converted our structured tables into OMOP format following OMOP CDM version 5.4.^26^. We manually mapped semantic concepts to standard representations in the OMOP vocabularies; these mappings were then validated by two clinical experts. For those source concepts without an existing standard representation, such as reflectance measurements of skin tone at a particular location with a specific device, we created custom standard concepts that we will eventually contribute back to the OMOP vocabulary team for future uptake into the community-curated vocabularies. In total, we created 2,704 concepts to represent the various imaging-related elements specific to this study. With regard to the data model itself, this dataset includes rows in the following tables: PERSON, VISIT_OCCURENCE, MEASUREMENT, OBSERVATION, DEVICE_EXPOSURE, PROCEDURE_OCCURRENCE, OBSERVATION_PERIOD, and CDM_SOURCE. A comprehensive summarization of the data flow can be found a Figure 1.

**Figure 1.**
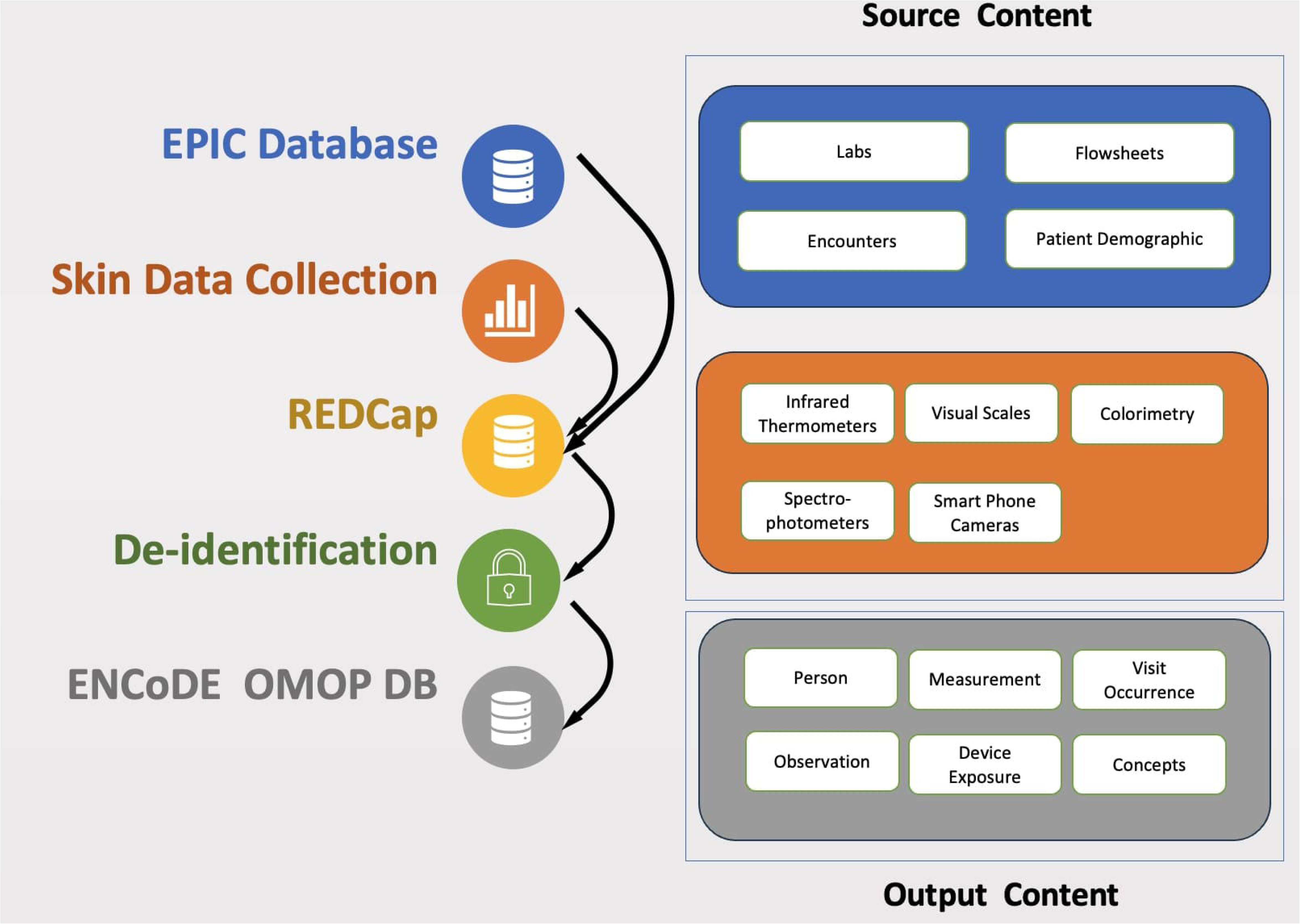
Data flow diagram and content. The left side of the figure represents how data flows in the collection process. Firstly, EHR data are pulled from EPIC databases into REDCap, and patient skin data is collected at the bedside and stored in REDCap. Then, the data are de-identified before leaving Duke’s compute enclave PACE via an honest broker request. Lately, data has been transformed into an OMOP format. The right side of the figures provides a high-level view of the data content. Source content contains patient’ EHR tables and data from five different types of devices or collection methods. Output content contains tables and images in OMOP format.

**Figure 2.**
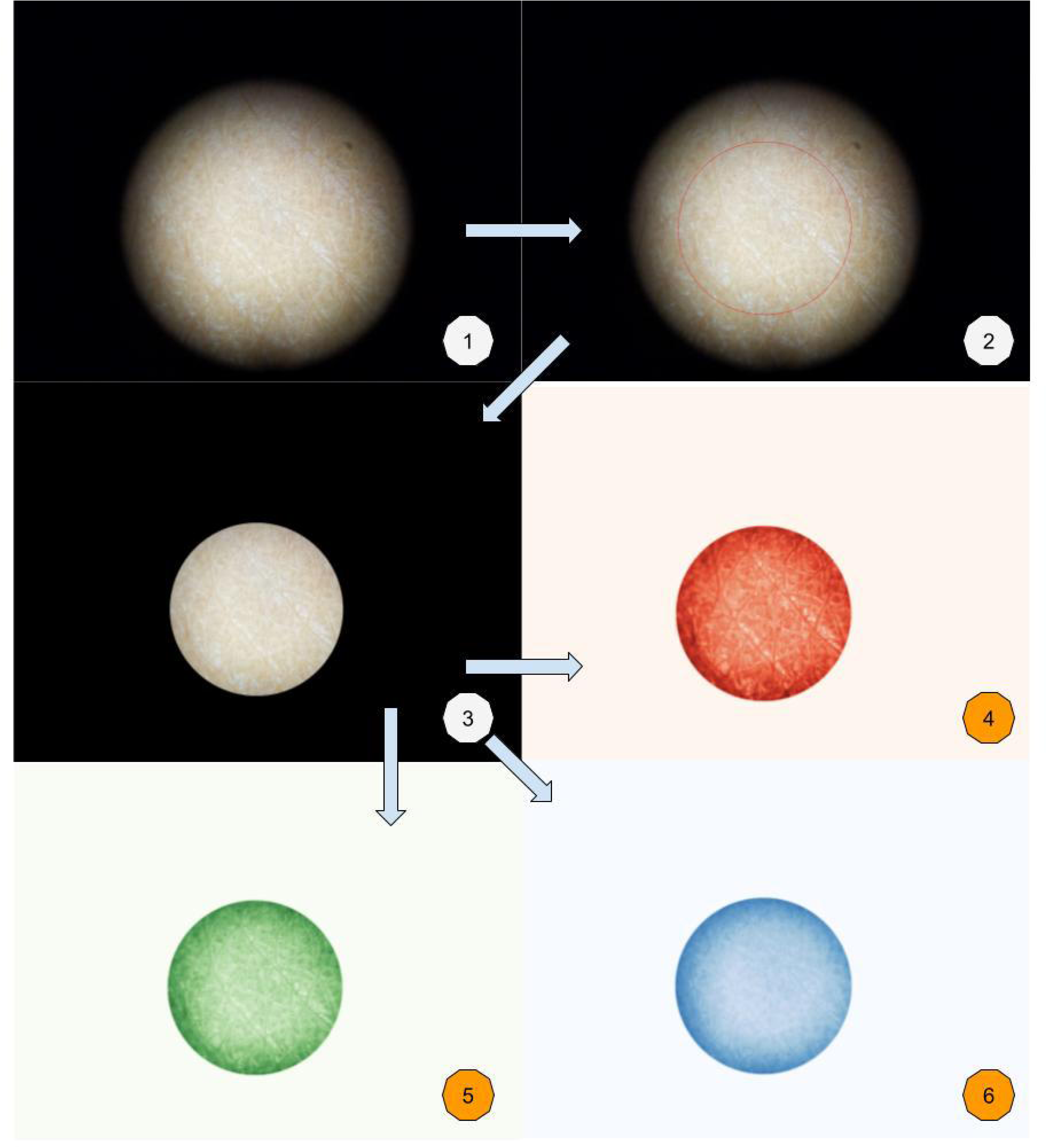
Image processing. This figure demonstrated how raw images were processed. (1) is the raw image taken with smartphone cameras. (2) The circle was calculated based on brightness represents the center of the image. (3) is the image output to the dataset, information inside the circle from (2) are kept. (4)-(6) are representations of how to derive image figures such as average red, green, and blue from the output image (3).

## Results

### Cohort

From January 2023 to June 2023, a total of 1,119 admitted inpatients with qualifying SaO_2_-SpO_2_ pairs were screened at Duke University Hospital. Out of those patients, 302 met our inclusion criteria and were approached, of whom 134 consented to this study. After six exclusions due to withdrawal or missing skin tone data, 128 patients were included in the final cohort. (39.8% female, 43% Black)

### Skin tone features

A total of 167 skin tone features from three administered visual scales, a colorimetry device (Delfin Technologies, SkinColorCatch), two spectrophotometer devices (Konica Minolta CM700d; Variable Inc, Spectro 1 Pro), and processed features from two types of mobile phone cameras (iPhone SE, hereafter referred to as iPhone; Google Pixel 4a, hereafter referred to as Android) were collected. As a medical concept, skin tone measurements have yet to be extensively investigated, and standardized concepts for skin tone at various locations are currently absent in the OMOP vocabulary. Therefore, skin tone concepts were created for this study; all the skin tone and skin temperature measurements were transformed into a long format and incorporated into the OMOP “measurement” table. These novel concepts were captured as OMOP’s “unit_concept_id” with skin tone at a specific location as “measurement_concept_id.” (e.g., measurement: unit:) Additionally, skin temperature was measured at the same locations using one clinical-range [clinical-range, 34-42.9 C°] temperature measurement device and one general-range [general-range, -32 - 500 C°] temperature measurement device. Figure 3 visualizes a few selected clinical features for one hospitalized patient

**Figure 3.**
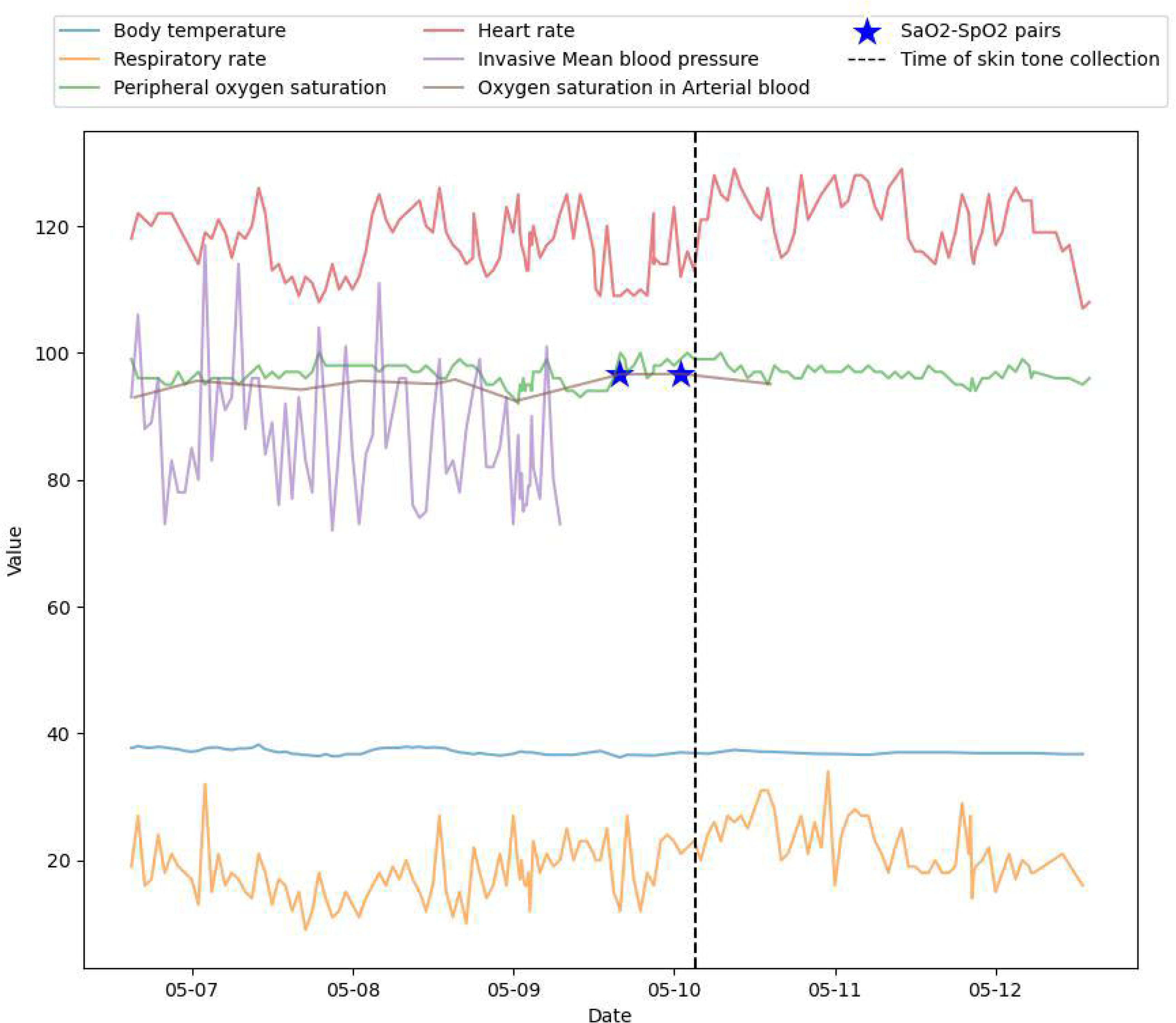
Sample data for a single patient. This is a timeline plot of a single patient’s data selected at random. The gold star represents the SaO2 - SpO2 pair we collected before skin data collection. The dashed black line represents the beginning of skin data collection. EHR data are available before and after skin collection.

### Cell phone images

To prevent the potential leak of Protected Health Information (PHI) for biometric imaging data, we released processed cell phone image data from 10 out of 16 body locations, excluding any territory containing finger, palm, or toe prints. With images missing due to other reasons described below, the open-source data files contain 1227 Android images and 1211 iPhone images. Images from biometric locations may be processed by the investigative team with code sharing and after IRB consent.

### Missing data

Missing data occurred occasionally in the cell phone data collection and two spectrophotometer devices due to technical issues or patient refusal. Twenty-nine patients are missing Variable Spectro 1 Pro measurements, and eight patients are missing Konica Minolta CM700d measurements. Detailed missingness rates for skin tone measurements can be found in Table 1. In the merged EHR clinical data, missingness occurred in vital signs and laboratory test values when no value was found within the set windows.

**Table 1.**
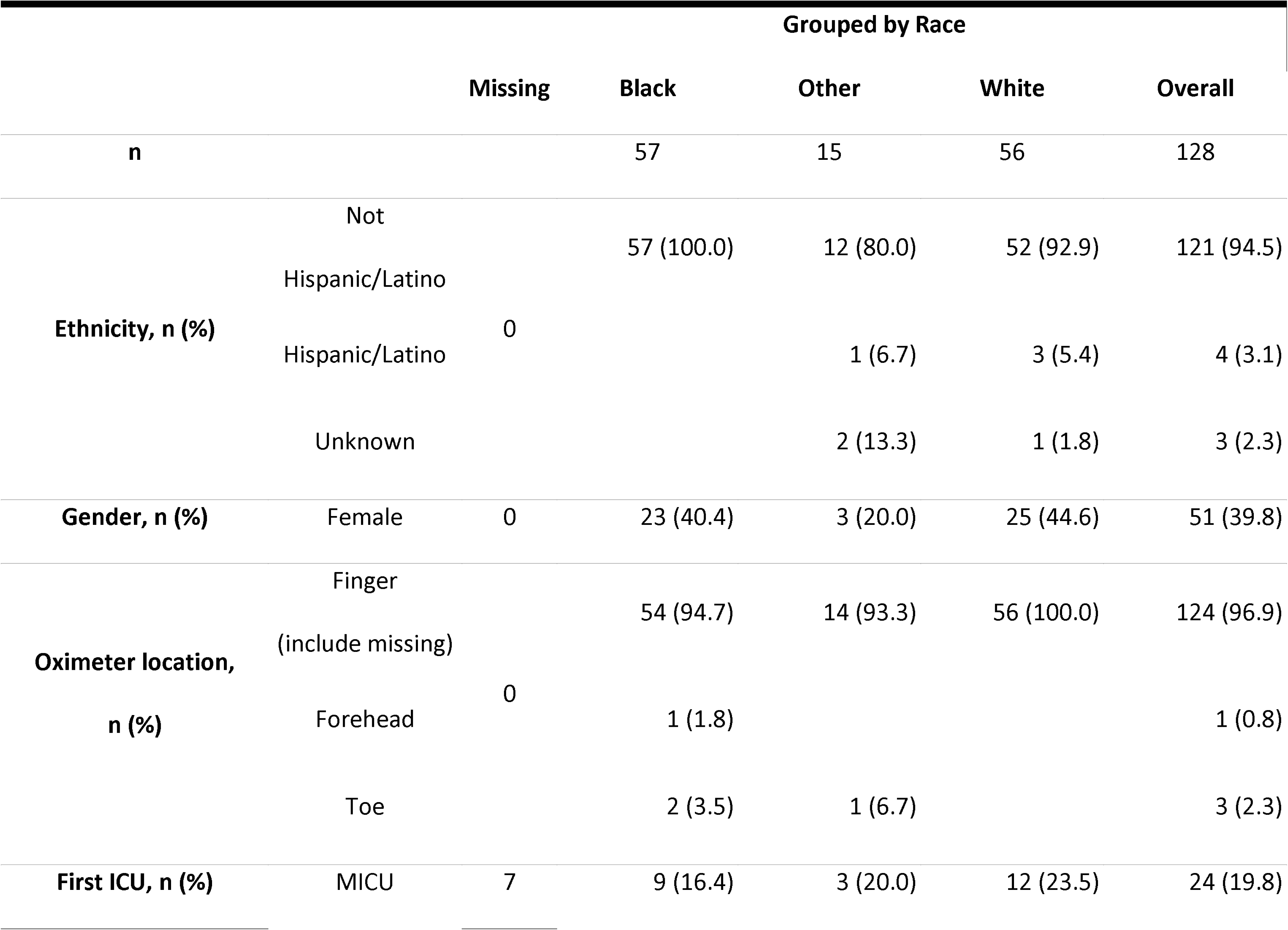
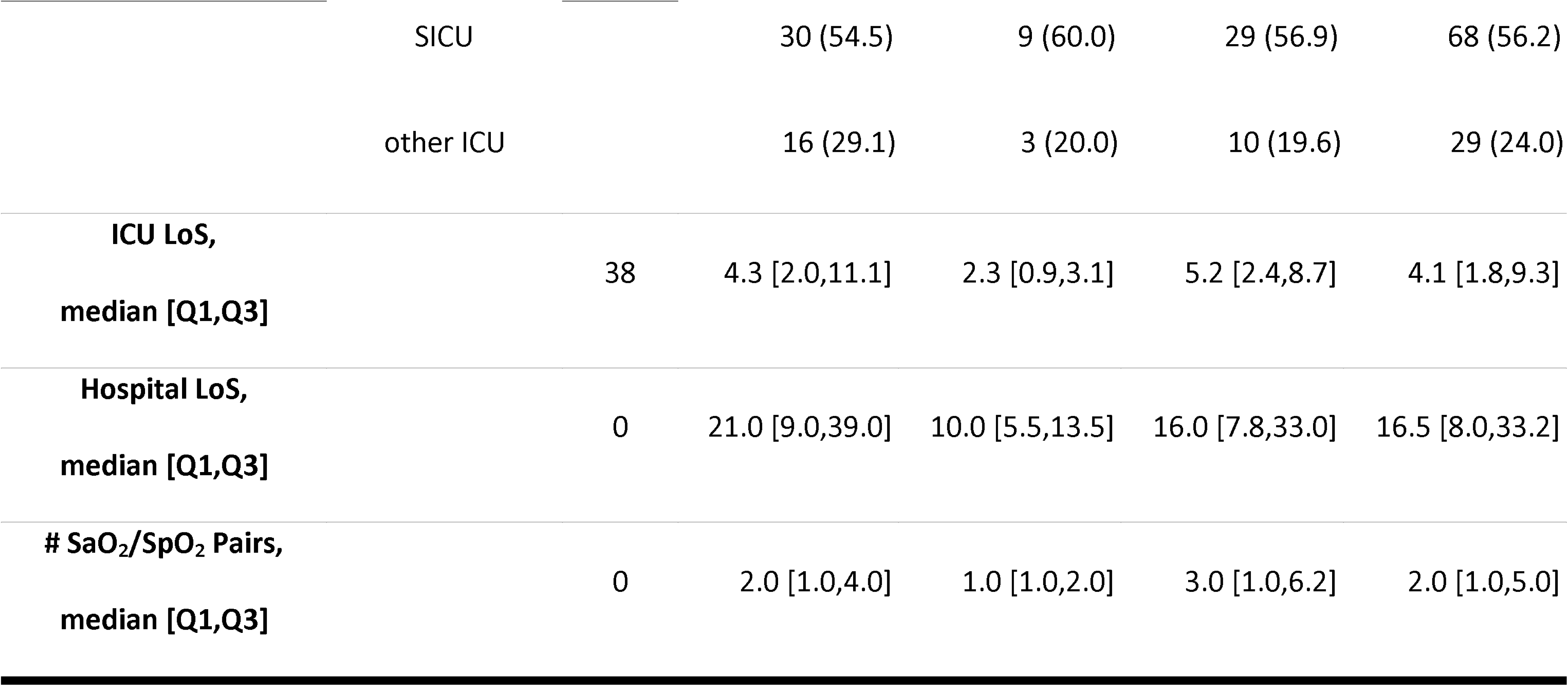
Characteristics of the study cohort. Demographic information for all 128 patients, along with their skin tone measurements, were grouped by race. The group “Other” contains patients who self-identify as Asian (n= 5), American Indian / Alaskan natives (n= 6), More than two races (n= 2), and Unknown race (n= 2).

**Table 2.**
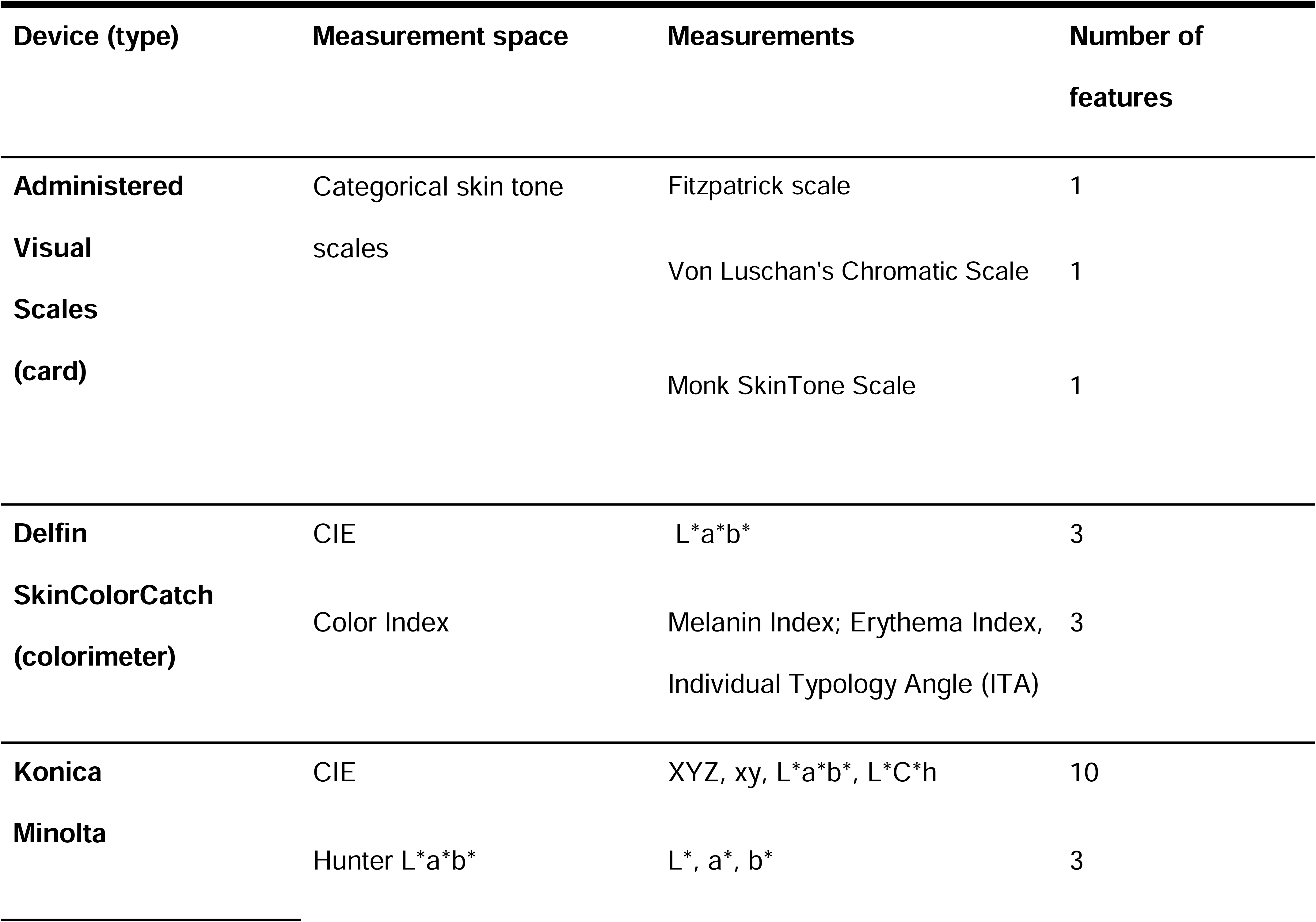
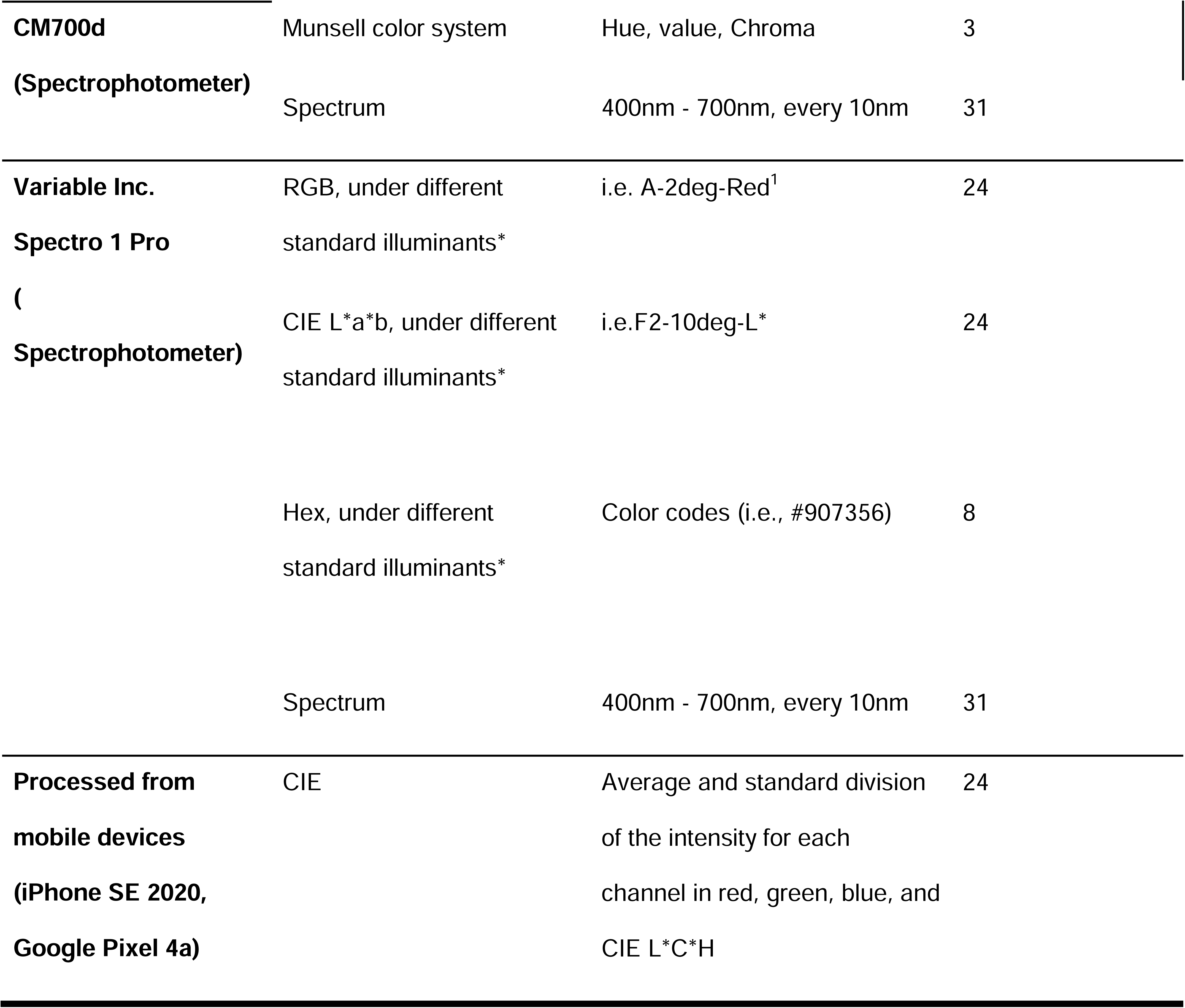
Structured skin tone features. This table describes all the features of skin tone measurement we collected in our dataset. Measurement devices and methods include administered visual scales (Fitzpatrick Skin Type, Monk Skin Tone, and Von Luschan); reflectance colorimetry (Delfin SkinColorCatch, Kuopio, Finland); and reflectance spectrophotometry (Variable Spectro 1 Pro Bridge Set, Variable, Inc TN, USA. and Konica Minolta CM-700D Spectrophotometer, Tokyo, Japan). In “A-2deg-Red”, “A” is the white point of standard illuminants, “2deg” is the field of view, which is short for 2 degrees, and “Red” is the color representation.

## Discussion

The ENCoDE project linked patients’ skin tone measurements across different body locations and patients’ EHR data. It pioneers the evaluation of skin tone as a medical concept to enhance health disparity research. Precise skin tone measurement can help identify patients’ biological skin color independently of their self-identified race. Our project is the first open-source dataset connecting skin tone from various measurement devices and methods on different body locations with 128 patients’ EHR data. This includes 167 skin tone features per location from multiple devices across 16 body locations, enabling the investigation of skin-associated health disparities with real-world clinical evidence. The dataset also includes images from smartphone devices, which allows for exploration of the feasibility of cell phone data collection as this can scale for cost-effectiveness. This project has the potential to develop more equitable AI tools to address biases and disparities associated with patients’ skin tone on medical devices. Such measurements promote diversity, equity, and inclusivity, contributing to advancements in personalized care.

By connecting skin tone measurements with patient EHR data, potential opportunities arise for researchers to investigate healthcare disparities associated with skin tone beyond pulse oximeter bias, such as temporal temperature measurement. ^27^ We mapped our dataset to the OMOP CDM for easy integration with other data sources. Due to the limited funding and resources, we couldn’t run this study in a multi-center setting with a larger population. Our vision is to create a community that can participate in providing feedback and create opportunities for collecting a multi-center dataset. Having a standard for collecting this new data is critical. This is why we chose a long format for all of our structured data and converted it into the OMOP format for a simplified, broader integration with other databases. The OMOP CDM standardizes healthcare data representation across diverse systems, enabling large-scale analytics and research. It has proven effective in converting from other data formats, such as MIMIC-IV. ^28,29^

In our dataset, the skin tone features from different measurement tools are measured together. The tools range from sophisticated but not commonly available spectrophotometers to widely used mobile phones and inexpensive print cards. However, in the most recent FDA guidelines, only the Monk SkinTone Scale (MTS) and Individual Typology Angle (ITA) are evaluated for capturing skin tone as a medical concept. ^22^ We measured over 167 skin tone features on different skin locations and put them side by side, creating opportunities for a thorough investigation of the advantages and disadvantages of various methods for measuring patients’ skin tone.

Additionally, we also included smartphone photos measured in a controlled lighting environment. As one of the most accessible devices, smartphones have great potential to be used as a tool for AI healthcare applications. ^30,31^ The advantage of smartphone images as a measurement for skin tone is the broad availabilities of the device as well as the abundance of datasets currently available. However, most current datasets focus on facial skin instead of skin location for medical applications such as fingers and palms. With the image data collected by smartphones across different body locations made available and linked with skin tone measurements from multiple scales and devices, potential work can create or validate equitable AI tools that utilize smartphone image data as affordable devices.

One limitation of our study is the relatively small number of patients from a single medical center due to limited resources. In the future, we aim to expand this dataset to include a larger sample size. Another limitation is the need for repeated measurements at the same scale and location. For administered scales, the measurer’s interpretation might have resulted in bias in the dataset. This design choice was made to maximize the patient sample size, given the constraints of staff and resources. In the future, we also seek to study the effect of the measurer’s race and sex on objective skin color measurements. Repeated measurements could take multiple days to complete, likely resulting in significant missing data due to the unpredictability of the clinical environment. A separate reliability experiment was conducted with a single patient across multiple days, which suggests inter-rater measurement variability for patient skin tone was much less than inter-patient variation in skin tone, even for the less precise administered visual scales. ^15^ Another limitation is that our skin color data are measured with specific lighting that might not be reproducible in other environments. This is mitigated using devices that directly provide controlled illumination (e.g., spectrophotometer and colorimeter, and to a lesser extent, mobile phones).

## Conclusions

The ENCoDE project provides the first openly shared dataset focused on skin tone measurements and linked to patients’ EHR data to study pulse oximetry bias. One hundred fifty-five skin tone features are collected, measuring different aspects of skin tone for each of the 16 body locations. Cell phone images for non-biometric body locations are collected further to investigate the relationship between images and curated features. The dataset is mapped to the OMOP Common Data Model, allowing ease of reusability and harmonization for future multi-center datasets. This dataset could potentially assist the collaboration between medical researchers and the medical AI communities to identify and combat skin tone-associated disparities, as well as provide more exploration that can guide regulatory bodies in evaluating pulse oximetry devices.

## Data Availability

This dataset is currently under review on PhysioNet.

